# Non-modifiable factors as moderators of the relationship between physical activity and brain volume: A cross-sectional UK Biobank study

**DOI:** 10.1101/2022.01.01.22268616

**Authors:** Belinda M Brown, Jaisalmer de Frutos Lucas, Tenielle Porter, Natalie Frost, Michael Vacher, Jeremiah J Peiffer, Simon M Laws

**Affiliations:** Centre for Healthy Ageing, Health Futures Institute, Murdoch University, Murdoch, Western Australia, Australia; Australian Alzheimer’s Research Foundation, Sarich Neuroscience Research Institute, Nedlands, Western Australia, Australia; Centre for Precision Health, Edith Cowan University, Joondalup, 6027, Western Australia, Australia; Laboratory of Cognitive and Computational Neuroscience, Center for Biomedical Technology, UPM-UCM, 28223 Pozuelo de Alarcón, Spain; Collaborative Genomics and Translation Group, School of Medical and Health Sciences, Edith Cowan University, Joondalup, 6027, Western Australia, Australia; School of Pharmacy and Biomedical Sciences, Faculty of Health Sciences, Curtin Health Innovation Research Institute, Curtin University, Bentley, Australia; Australian e-Health Research Centre, CSIRO, Floreat 6014, Western Australia, Australia

## Abstract

**Background:** Grey matter atrophy occurs as a function of ageing and is accelerated in dementia. Previous research suggests physical activity attenuates grey matter loss; however, there appears to be individual variability in this effect. Understanding factors that can affect the relationship between physical activity and brain volume may enable prediction of individual response, and aid in identifying those that gain the greatest neural benefits from physical activity. The current study examined the relationship between objectively-measured physical activity and brain volume; and whether this relationship is moderated by age, sex, or *a priori* candidate genetic factors.

**Methods:** Data from 10,083 men and women (50 years and over) of the UK Biobank were used to examine: 1) the relationship between objectively-measured physical activity and brain volume; and 2) whether the relationship between objectively-measured physical activity and brain volume is moderated by age, sex, brain-derived neurotrophic factor (*BDNF*) Val66Met, or apolipoprotein (*APOE*) ε4 allele carriage. All participants underwent a magnetic resonance imaging scan to quantify grey matter volumes, physical activity monitoring via accelerometry, and genotyping.

**Results:** Physical activity was associated with total grey matter volume (B = 0.14, *p* = 0.001, *q* = 0.005) and right hippocampal volume (B = 1.45, *p* = 0.008, *q* = 0.016). The physical activity*sex interaction predicted cortical grey matter (B = 0.22, *p* = 0.003, *q* = 0.004), total grey matter (B = 0.30, *p* < 0.001, *q* = 0.001), and right hippocampal volume (B = 3.60, *p* = 0.001, *q* = 0.002). Post-hoc analyses revealed males received benefit from higher physical activity levels, in terms of greater cortical grey matter volume (B = 0.13, *p* = 0.01), total grey matter volume (B=0.23, *p* < 0.001), and right hippocampal volume (B = 3.05, *p* = 0.008). No moderating effects of age, *APOE* ε4 allele carriage, or *BDNF* Val66Met genotype were observed.

**Discussion:** Our results indicate that in males, but not females, an association exists between objectively-measured physical activity and grey matter volume. Future research should evaluate longitudinal brain volumetrics to better understand the nature of sex-effects on the relationship between physical activity and brain volume.

## INTRODUCTION

Atrophy of grey matter occurs as a function of ageing and is accelerated in neurodegenerative conditions, such as dementia^1^. There is no effective pharmaceutical treatment for dementia or cognitive decline that alters the course of disease progression, nor that has been demonstrated to attenuate grey matter loss associated with either ageing or disease. Recent updates from the Lancet Commission on Dementia Prevention^2^ posits nine modifiable risk factors as contributing to dementia risk, one of which is physical inactivity. Indeed, higher levels of habitual physical activity and exercise interventions have both been shown to either attenuate age-related grey matter volume loss^3–5^, or in the case of some brain regions (i.e. the hippocampus; brain region involved in learning and memory) contribute to increases in grey matter volume^6–9^. Nevertheless, there appears to be some distinct heterogeneity in study findings across this field^10–12^, and individual variability in response likely influences the relationship between physical activity and brain volume^13^.

Understanding moderators of the relationship between physical activity and brain volume is vital, as it may enable the identification of individuals, or groups of individuals, that gain the greatest neural benefits from physical activity^13, 14^. A moderating variable is one that changes the effect of an exposure on an outcome, i.e., a variable that influences the relationship between physical activity and brain volume. Moderation is confirmed when the relationship between two variables is different across various levels of the moderating variable. In the case of physical activity and brain volume, a number of potential moderators have been posited and investigated, including non-modifiable between-person factors such as age, sex, and genetics.

Age may influence the relationship between physical activity and brain volume, whereby an effect may only be discernible once reaching, or within, a particular age range. For example, age may influence the effects of physical activity on important mediating molecules, such as brain-derived neurotrophic factor (BDNF)^15^. Sex has also emerged as a possible moderator of the relationship between physical activity and brain health. Females tend to be disproportionately affected by Alzheimer’s disease, with females observed to have higher rates of incidence, greater accumulation of associated pathologies, and greater decreases in brain volume^16^. Based on sex differences in physiological response to physical activity^17, 18^, it is possible the beneficial effects of physical activity on the brain may differ between males and females. Literature examining the relationship between physical activity and cognition indicates females typically gain more benefits from higher levels of activity^19^. Nevertheless, the examination of sex-effects in the relationships between physical activity and brain volume have not been as consistent, with some studies identifying only males^20^ and others only females^21, 22^ gain benefit from higher physical activity levels.

A substantial literature exists to suggest that salient genetic factors play a role in moderating the relationship between physical activity and brain health. One such potential moderating factor is a single nucleotide polymorphism of the *BDNF* gene, termed Val66Met, which influences the secretion of BDNF: a potent neuromodulator, associated with better brain health and the levels of which are decreased in Alzheimer’s disease. Given the independent link between *BDNF* Val66Met carriage and smaller brain volume^23, 24^, and the potent effect of exercise in increasing circulating BDNF levels^25^, there is a strong theoretical basis to suggest a moderating effect of this genetic factor. Indeed, one report suggests Val/Val homozygotes, but not Met carriers, have a relationship between higher levels of physical activity and brain volume^26^. The apolipoprotein (*APOE*) ε4 allele is the greatest known genetic risk factor for sporadic Alzheimer’s disease and is also associated with swifter cognitive decline and smaller brain volume in cognitively normal older adults. To our knowledge only one study (*n* = 97) has examined the moderating effects of *APOE* ε4 carriage on the relationship between physical activity and brain volume, finding that only ε4 carriers had reduced hippocampal atrophy in relation to higher physical activity levels^4^.

Previous studies of moderators of the physical activity and brain volume relationship have examined non-modifiable moderators in isolation, and/or in relatively small samples. The current study will build on prior work by utilising data from a large cohort study (UK Biobank) to examine the moderating effects of a number of non-modifiable factors that may influence the relationship between physical activity and brain volume. We will address two specific aims: 1) Examine the relationship between objectively-measured physical activity and brain volume in the UK Biobank; and 2) Examine whether the relationship between objectively-measured physical activity and brain volume is moderated by age, sex, *BDNF* Val66Met, or *APOE* ε4 allele carriage.

## METHODS

### Participants

The UK Biobank (http://ukbiobank.ac.uk/) is a large prospective cohort study that commenced in 2006 and has collected health data on over 500,000 participants^27^. Individuals aged 40-69 years were recruited to attend one of 22 data collection sites between 2006 and 2010 for a baseline assessment. Data pertinent to the current study was collected in 2013-2015 (accelerometer data) and 2014 onward (magnetic resonance imaging; MRI, scans).

### Standard Protocol Approvals, Registrations, and Consents

Ethical approval was provided by the NHS National Research Ethics Committee (11/NW/0382). The current analyses were conducted as a part of the UK Biobank application 45567. All participants provided informed consent prior to participation.

### Exclusion and inclusion criteria

The dataset received from UK Biobank contained data from 502,527 participants (eFigure 1). As our research focus is understanding the relationship between physical activity and brain volume in older adults, we only included individuals aged 50 years and older at baseline (*n* = 384,648). In addition, we excluded participants with a diagnosis of Alzheimer’s disease, dementia (of any kind), or mild cognitive disorder (*n* = 1917). Finally, only participants with complete accelerometer and volumetric MRI data as well as data required for covariates (outlined in further detail below) were used in the analyses presented here (*n* = 10,083).

**Figure 1:**
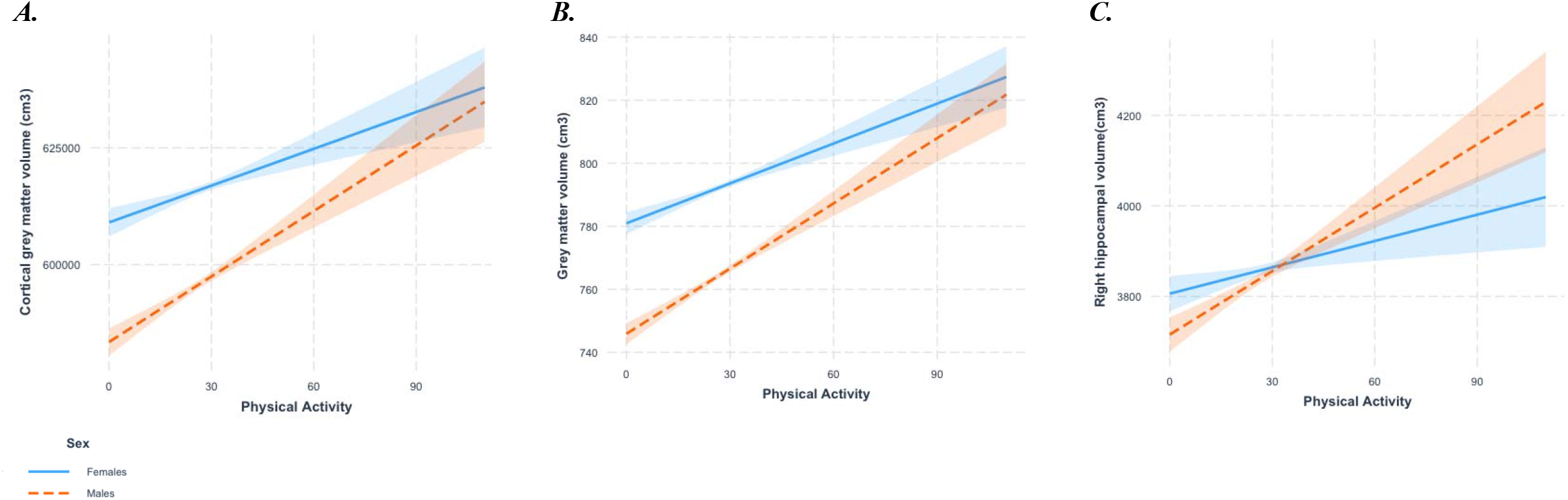
Interaction between physical activity (adjusted average daily accelerations) and sex on A) Cortical grey matter volume (normalised for head size scaling factor); B) Grey matter volume (normalised for head size scaling factor); C) Right hippocampal volume (normalised for head size scaling factor). Dotted orange line represents males and solid blue line represents females.

### Habitual physical activity measurement

Between February 2013 and December 2015, participants of the UK Biobank were invited at random to undergo objective habitual physical activity assessment through wearing an accelerometer for 7 days. From June 2013, participants that agreed to participate were sent Axivity AX3 wrist-worn triaxial accelerometers. The accelerometer units were programmed to commence data collection at 10am two working days after postage and continue to collect acceleration data during wear-time on the dominant wrist over the following 7 days at 100Hz with a dynamic range of ± 8g. Within the current study, we have used average daily acceleration, adjusted for wear time (in days), as our measure of habitual physical activity.

### Structural magnetic resonance imaging

Volumetric MRI data were acquired using a Siemens Skyra 3T scanner (Siemens Healthcare, Erlangen, Germany), with a standard 32-channel RF receive head coil as per procedures outlined by the UK Biobank^28^. Briefly, the 3D MPRAGE T1-weighted images underwent acquired pre-processing and analysis using FSL packages (version 5, FMRIB Software Library, Oxford, England). Imaging-derived phenotypes, specifically grey matter volumes, were generated by the image-processing pipeline developed by, and run on behalf of, the UK Biobank^29^. All brain volume variables used within the current study have been corrected for head size.

### Genotyping

Genotying for the UK Biobank was conducted by Affymetrix on purpose-designed arrays; ~50,000 samples were analysed using the BiLEVE Axiom array and ~450,000 on the Affymetric UK Biobank Axiom array. Details of quality control practices have been described previously^30^. *BDNF*Val66Met polymorphism was used to classify participants as either Met carriers or Val/Val homozygotes. In addition, participants were identified as either *APOE* ε4 allele carriers or non-carriers.

### Demographic and medical data

A number of variables were identified as potential covariates in the relationship between physical activity and brain volume, including education, depression, socioeconomic status, overall health rating, and vascular conditions and diabetes.

Participants reported their highest educational attainment (seven categories provided) and socioeconomic status was estimated using the Townsend deprivation index^31^. Participants were asked whether their doctor had diagnosed them with diabetes, hypertension, myocardial infarction, angina, stroke or hypertension. Overall health was self-reported and scored on a 4-point scale from ‘poor’ to ‘excellent’. For any of the above variables, participants that stated ‘prefer not to answer’ or with missing data, were excluded from the analyses.

### Statistical analysis

Statistical analyses were conducted in R (R Core Team; 2013) and Statistical Package for the Social Sciences (SPSS, IBM, Version 24). P-values from independent variables of interest and interactions were corrected for multiple comparisons using false discovery rate^32^. Descriptive statistics were calculated for demographic and outcome variables, and differences across sex, *APOE* ε4 allele and *BDNF* Val66Met polymorphism carriage were examined using independent samples t-tests (continuous variables) and chi-square analyses (categorical variables).

Linear models were used to examine the relationship between habitual objectively-measured physical activity (average daily accelerations; independent variable) and brain volume (dependent variable) in all participants, adjusting for potential confounders: age, sex, body mass index, *APOE* ε4 allele, *BDNF* Val66Met, education, depression, Townsend deprivation index, overall health rating, and doctor diagnosed vascular conditions and diabetes (‘A’ models). The interaction terms physical activity*age, physical activity*sex, physical activity\**APOE*, physical activity\**BDNF* were then added to ‘A’ models to examine the moderating effect of non-modifiable factors on the relationship between physical activity and brain volume (‘B’ models).

## RESULTS

Examination of health and demographic differences between sexes (Table 1) revealed males were older (*t* = 11.0, *p* < 0.001), had a higher body mass index (*t* = 11.1, *p <* 0.001), lower levels of objectively-measured habitual physical activity (*t* = 6.6, *p* < 0.001), poorer self-reported health ratings (χ^2^ = 15.4, *p* < 0.01), higher prevalence of vascular conditions (χ^2^ = 143.5, *p* < 0.001) and diabetes (χ^2^ = 25.0, *p* < 0.001), compared with female participants. In addition, cross-sectional evaluations of normalised brain volume differences revealed males had smaller volumes of total grey matter (*t* = 34.5, *p* < 0.001), cortical grey matter (*t* = 28.6, *p* < 0.001), and right hippocampal volume (*t* = 2.1, *p* < 0.05; analysis not corrected for age or other covariates). Health and demographic details across *APOE* ε4 and *BDNF* Val66Met are detailed in eTable 1. The genotypic distribution of all polymorphisms of interest did not deviate from Hardy-Weinberg Equilibrium (rs6265, *p* = 0.20; rs429358, *p* = 0.43; rs7412, *p* = 0.95).

**Table 1:** Descriptive statistics of UK Biobank participants at baseline (2006-2010) aged over 50 with accelerometer and MRI data, stratified by sex.

|  | **Females**<br>(*n* = 5396) | **Males**<br>(*n* = 4687) | **Test statistic** |
| --- | --- | --- | --- |
| **Age (years)** | 58.2 ± 4.9 | 59.3 ± 5.2 | *t* = 11.0*** |
| **Body mass index** | 26.0 ± 4.4 | 26.9 ± 3.7 | *t* = -11.1*** |
| **Physical activity (accelerometer daily accelerations)** | 28.0 ± 8.3 | 26.9 ± 9.0 | *t* = 6.6*** |
| **Accelerometer wear time (days)** | 6.4 ± 1.3 | 6.4 ± 1.4 | *t* = 0.56 |
| **Education** |  |  |  |
| College or University degree, % (*n*) | 43.6 (2355) | 48.1 (2255) | $\chi^2 = 169.8^{***}$ |
| A levels/AS levels or equivalent, % (*n*) | 13.6 (736) | 11.9 (557) |  |
| O levels/DCSEs or equivalent, % (*n*) | 21.4 (1156) | 16.6 (780) |  |
| CSEs or equivalent, % (*n*) | 3.4 (181) | 3.0 (139) |  |
| NVQ/HND/HNC or equivalent, % (*n*) | 3.3 (180) | 8.1 (381) |  |
| Other professional qualification, % (*n*) | 6.9 (375) | 4.9 (229) |  |
| None of the above, % (*n*) | 7.6 (413) | 7.4 (346) |  |
| **APOE ε4 carriage % (*n*)** | 27.7 (1493) | 26.9 (1261) | $\chi^2 = 0.74$ |
| **BDNF Val66Met carrier % (*n*)** | 34.6 (1867) | 34.3 (1607) | $\chi^2 = 0.11$ |
| **Major depression % (*n*)** | 21.7 (1169) | 21.1 (991) | $\chi^2 = 0.40$ |
| **Townsend Deprivation Index** | -2.0 ± 2.6 | -2.1 ± 2.6 | *t* = 1.5 |
| **Overall health rating** |  |  |  |
| Excellent, % (*n*) | 26.4 (1427) | 25.1 (1177) | $\chi^2 = 15.4^{**}$ |
| Good, % (*n*) | 60.9 (3286) | 59.8 (2802) |  |
| Fair, % (*n*) | 11.2 (603) | 13.7 (640) |  |
| Poor, % (*n*) | 1.4 (74) | 1.4 (65) |  |
| **Vascular problems % (*n*)** | 10.9 (591) | 18.0 (843) | $\chi^2 = 143.5^{***}$ |
| **Diabetes** | 1.8 (96) | 3.3 (155) | $\chi^2 = 25.0^{***}$ |
| **Head scaling factor** | 1.4 ± 0.1 | 1.2 ± 0.1 | *t* = 78.1*** |
| **Cortical grey matter (normalised; cm<sup>3</sup>)** | 615.9 ± 35.6 | 595.7 ± 35.3 | *t* = 28.6*** |
| **Grey matter (normalised; cm<sup>3</sup>)** | 792.3 ± 40.4 | 764.1 ± 41.2 | *t* = 34.5*** |
| **Left hippocampal volume (normalised; mm<sup>3</sup>)** | 3748.3 ± 419.5 | 3732.1 ± 489.3 | *t* = 1.8 |
| **Right hippocampal volume (normalised; mm<sup>3</sup>)** | 3857.0 ± 417.6 | 3837.1 ± 514.2 | *t* = 2.1* |

\**p* < 0.05, \*\**p* < 0.01; \*\*\**p* < 0.001; Chi-square analyses for categorical variables and independent sample t-tests were conducted for continuous variables. Abbreviations: APOE,
apolipoprotein E; *BDNF* Val66Met, brain-derived neurotrophic factor Valine66Methionine single nucleotide polymorphism.

Physical activity was associated with larger total grey matter volume (B = 0.14, *p* = 0.001, *q* = 0.005) and right hippocampal volume (B = 1.45, *p* = 0.008, *q* = 0.016; Table 3 & 5). When examining factors that may affect the relationship between physical activity and brain volume, only sex was found to have a moderating effect. The physical activity*sex interaction predicted cortical grey matter (B = 0.22, *p* = 0.003, *q* = 0.004), total grey matter (B = 0.30, *p* < 0.001, *q* = 0.001) and right hippocampal volume (B = 3.60, *p* = 0.001, *q* = 0.002; Tables 2–5). Post-hoc analyses, following stratification of the cohort by sex, revealed that only males had an association between physical activity levels and greater cortical grey matter volume (B = 0.13, *p* = 0.01), total grey matter volume (B = 0.23, *p* < 0.001), and right hippocampal volume (B = 3.05, *p* < 0.001; Figures 1A-C). Neither the physical activity*age interaction nor either of physical activity\**APOE* or physical activity\**BDNF* interactions were significant in predicting any brain volume measure.

**Table 2:** The relationship between habitual physical activity levels (average acceleration adjusted for time worn) and cortical grey matter volume (normalised for head size scaling factor) from magnetic resonance imaging

| DV: Cortical grey matter volume (cm<sup>3</sup>) | Model 1A |  |  | Model 1B |  |  |
| --- | --- | --- | --- | --- | --- | --- |
|  | Estimate (B) | LL 95% CI | UL 95% CI | Estimate (B) | LL 95% CI | UL 95% CI |
| PA | 0.07 | 0.00 | 0.15 | -0.33 | -1.16 | 0.50 |
| Age | -3.08*** | -3.21 | -2.96 | -3.22*** | -3.62 | -2.81 |
| Sex | -16.37*** | -17.63 | -15.11 | -22.31*** | -26.47 | -18.15 |
| BMI | -0.22** | -0.38 | -0.06 | -0.23** | -0.39 | -0.07 |
| Assessment centre | 0.55*** | 0.43 | 0.67 | 0.55*** | 0.43 | 0.67 |
| APOE ε4 allele carriage | -0.59 | -1.98 | 0.80 | -2.01 | -6.65 | 2.62 |
| BDNF Val66Met | -0.07 | -1.37 | 1.24 | 0.45 | -3.91 | 4.80 |
| Education | -0.21 | -0.43 | 0.02 | -0.21 | -0.43 | 0.02 |
| Depression | 2.58** | 1.05 | 4.12 | 2.58** | 1.05 | 4.11 |
| Townsend Deprivation Index | -0.28* | -0.51 | -0.04 | -0.28* | -0.51 | -0.04 |
| Overall health rating | 0.13 | -0.78 | 1.05 | 0.12 | -0.80 | 1.04 |
| Diabetes | -6.34** | -10.30 | -2.38 | -6.13** | -10.09 | -2.16 |
| Vascular problems | -0.38 | -0.85 | 0.10 | -0.38 | -0.85 | 0.10 |
| PA x Age |  |  |  | 0.00 | -0.01 | 0.02 |
| PA x Sex |  |  |  | 0.22** | 0.07 | 0.36 |
| PA x APOE |  |  |  | 0.05 | -0.11 | 0.21 |
| PA x BDNF |  |  |  | -0.02 | -0.17 | 0.13 |
| F-statistic for linear model | | | $F_{13,10070} = 275.0^{***}$ | | | $F_{17,10066} = 211.1^{***}$ |

\* $p < 0.05$ ; \*\* $p < 0.01$ ; \*\*\* $p < 0.001$ . Abbreviations: 95% CI, 95% confidence interval; APOE ε4, apolipoprotein ε4 allele carriage; B, unstandardised beta value from linear model; BDNF Val66Met, brain-derived neurotrophic factor Valine66Methionine single nucleotide polymorphism; BMI, body mass index; LL, lower limit; UL, upper limit; PA, objectively-quantified physical activity (average accelerations adjusted for wear time).

**Table 3:** The relationship between habitual physical activity levels (average acceleration adjusted for time worn) and grey matter volume (normalised for head size scaling factor) from magnetic resonance imaging

| DV: Grey matter volume (cm<sup>3</sup>) | Model 2A |  |  | Model 2B |  |  |
| --- | --- | --- | --- | --- | --- | --- |
|  | Estimate (B) | LL 95% CI | UL 95% CI | Estimate (B) | LL 95% CI | UL 95% CI |
| PA | 0.14** | 0.05 | 0.22 | -0.49 | -1.42 | 0.45 |
| Age | -3.81*** | -3.95 | -3.67 | -4.03*** | -4.48 | -3.57 |
| Sex | -22.88*** | -24.29 | -21.46 | -31.04*** | -35.71 | -26.37 |
| BMI | -0.74*** | -0.92 | -0.55 | -0.75*** | -0.94 | -0.57 |
| Assessment centre | 0.56*** | 0.43 | 0.69 | 0.56*** | 0.43 | 0.69 |
| APOE ε4 allele carriage | -0.40 | -1.96 | 1.17 | -2.56 | -7.76 | 2.65 |
| BDNF Val66Met | -0.34 | -1.80 | 1.13 | -0.28 | -5.17 | 4.62 |
| Education | -0.20 | -0.45 | 0.05 | -0.20 | -0.45 | 0.05 |
| Depression | 2.02* | 0.30 | 3.75 | 2.02* | 0.30 | 3.75 |
| Townsend Deprivation Index | -0.42** | -0.69 | -0.15 | -0.42** | -0.69 | -0.15 |
| Overall health rating | 0.37 | -0.67 | 1.40 | 0.35 | -0.68 | 1.38 |
| Diabetes | -10.91*** | -15.36 | -6.46 | -10.63*** | -15.08 | -6.17 |
| Vascular problems | -0.33 | -0.86 | 0.21 | -0.33 | -0.86 | 0.21 |
| PA x Age |  |  |  | 0.01 | -0.01 | 0.02 |
| PA x Sex |  |  |  | 0.30*** | 0.14 | 0.46 |
| PA x APOE |  |  |  | 0.08 | -0.10 | 0.26 |
| PA x BDNF |  |  |  | 0.00 | -0.17 | 0.17 |
| F-statistic for linear model | | $F_{13,10070} = 365.0^{***}$ | | | $F_{17,10066} = 280.3^{***}$ | |

\* $p < 0.05$ ; \*\* $p < 0.01$ ; \*\*\* $p < 0.001$ . Abbreviations: 95% CI, 95% confidence interval; APOE ε4, apolipoprotein ε4 allele carriage; B, unstandardised beta value from linear model; BDNF Val66Met, brain-derived neurotrophic factor Valine66Methionine single nucleotide polymorphism; BMI, body mass index; LL, lower limit; UL, upper limit; PA, objectively-quantified physical activity (adjusted average accelerations).

**Table 4:** The relationship between habitual physical activity levels (average acceleration adjusted for time worn) and left hippocampal volume (normalised for head size scaling factor) from magnetic resonance imaging

| DV: Left hippocampal volume (mm<sup>3</sup>) | Model 3A |  |  | Model 3B |  |  |
| --- | --- | --- | --- | --- | --- | --- |
|  | Estimate (B) | LL 95% CI | UL 95% CI | Estimate (B) | LL 95% CI | UL 95% CI |
| PA | 0.96 | -0.08 | 1.99 | 0.22 | -11.28 | 11.72 |
| Age | -22.63*** | -24.36 | -20.90 | -22.71*** | -28.34 | -17.08 |
| Sex | 14.07 | -3.30 | 31.45 | -18.31 | -75.69 | 39.08 |
| BMI | -1.16 | -3.40 | 1.08 | -1.24 | -3.48 | 1.00 |
| Assessment centre | 3.59*** | 1.97 | 5.21 | 3.58*** | 1.96 | 5.19 |
| APOE ε4 allele carriage | -33.32** | -52.50 | -14.14 | -6.65 | -70.66 | 57.36 |
| BDNF Val66Met | 11.52 | -6.45 | 29.50 | -6.02 | -66.17 | 54.13 |
| Education | -0.14 | -3.20 | 2.93 | -0.13 | -3.19 | 2.93 |
| Depression | 18.79 | -2.37 | 39.96 | 18.98 | -2.20 | 40.15 |
| Townsend Deprivation Index | -5.25** | -8.53 | -1.96 | -5.25** | -8.53 | -1.96 |
| Overall health rating | -8.09 | -20.78 | 4.59 | -8.11 | -20.79 | 4.58 |
| Diabetes | -59.25* | -113.88 | -4.62 | -57.97* | -112.65 | -3.30 |
| Vascular problems | -9.29** | -15.84 | -2.74 | -9.28** | -15.83 | -2.73 |
| PA x Age |  |  |  | 0.00 | -0.19 | 0.20 |
| PA x Sex |  |  |  | 1.18 | -0.82 | 3.17 |
| PA x APOE |  |  |  | -0.96 | -3.18 | 1.25 |
| PA x BDNF |  |  |  | 0.63 | -1.45 | 2.72 |
| F-statistic for linear model | | $F_{13,10061} = 59.2^{***}$ | | | $F_{17,10057} = 45.4^{***}$ | |

\* $p < 0.05$ ; \*\* $p < 0.01$ ; \*\*\* $p < 0.001$ . Abbreviations: 95% CI, 95% confidence interval; APOE ε4, apolipoprotein ε4 allele carriage; B, unstandardised beta value from linear model; BDNF Val66Met, brain-derived neurotrophic factor Valine66Methionine single nucleotide polymorphism; BMI, body mass index; LL, lower limit; UL, upper limit; PA, objectively-quantified physical activity (adjusted average accelerations).

**Table 4:**
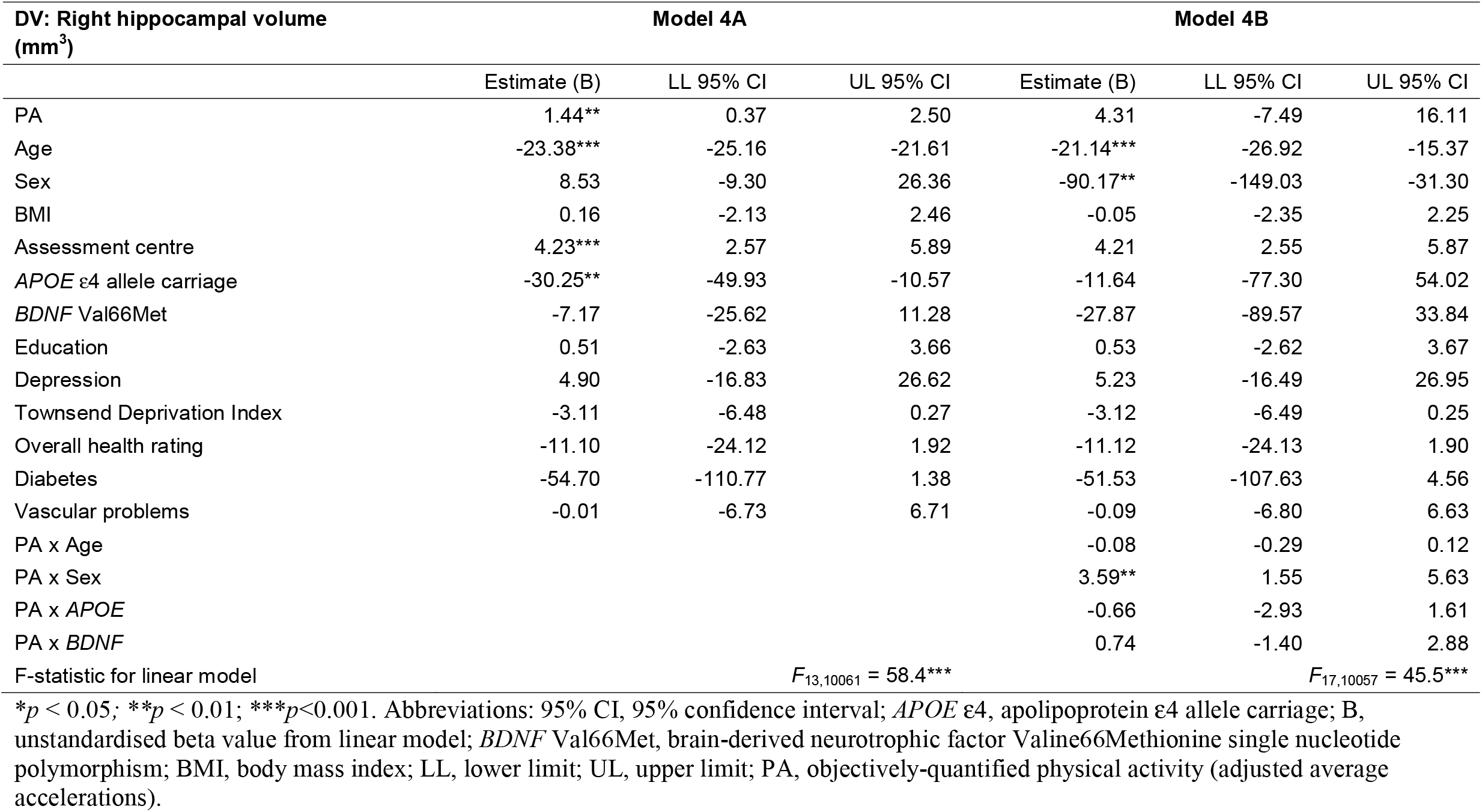
The relationship between habitual physical activity levels (average acceleration adjusted for time worn) and right hippocampal volume (normalised for head size scaling factor) from magnetic resonance imaging

## DISCUSSION

The aim of the current study was to examine non-modifiable factors as moderators of the relationship between habitual objectively-measured physical activity and brain volume in a large population-based cohort of older adults (aged 50 years and over). In our total dataset of 10,083 older men and women, we found objectively-measured physical activity levels were associated with larger total grey matter volume and right hippocampal volume. In addition, we observed that sex moderated the relationship between physical activity and cortical grey matter, total grey matter, and right hippocampal volume; whereby only males had an association between objectively-measured physical activity and these brain volume measurements. We did not observe any moderating effects of age, *APOE* ε4 allele carriage, or the *BDNF* Val66Met polymorphism on the relationship between physical activity and brain volume.

We report higher physical activity levels are linked with larger total grey matter and right hippocampal volumes in our large sample. Our findings build on earlier work using UK Biobank data that report objectively measured physical activity to be associated with total grey matter volume and both left and right hippocampal volumes in 5272 adults^33^. Our contrasting findings regarding left hippocampal volume could be attributed to differences in our samples, e.g., we implemented a minimum age cut-off of 50 years. Nevertheless, the literature regarding effects of physical activity and exercise on both right and left hippocampal volume is complex and inconsistent. Some studies report relationships between physical activity and both left and right hippocampal volumes^34^, some studies report effects on only the left^12^ and others only the right hippocampus^11^. The mechanisms underlying these differential effects remain to be elucidated; however, it is possible that physical activity-induced BDNF may act differently on the left and right hippocampi in different samples.

Future research should also consider examining the functions of the left and right hippocampus (verbal and visual-spatial memory respectively), as this may provide insight into the functional outcomes of the physical activity-hippocampus relationship^35^. Indeed, previous work in this field suggests that in natural environments, physical activity has been associated with increases in cognitive demands on spatial orientation and memory, which may underpin the links between physical activity and brain health, and more specifically right hippocampal volume^36^.

Our study revealed the relationship between physical activity and total grey matter and right hippocampal volumes was confined to males only. The current literature regarding sex differences in the physical activity-brain volume relationship is relatively small. Our findings are consistent with longitudinal work from Barha and colleagues^20^ who identified higher levels of self-reported walking at baseline was associated with larger hippocampal volume 10 years later in males only, and larger dorsolateral prefrontal cortex in females only. It is possible that different brain regions are affected differently by physical activity in males versus females: this is reflected by functional data suggesting females are more likely to gain benefits from physical activity in terms of executive function (i.e., frontal lobe mediated brain function)^37^. In addition, BDNF expression is reduced in post-menopausal women, and although this effect can be partially attenuated with structured exercise, females may not achieve the same neurotrophic response as males^38^. Nevertheless, in a smaller cross-sectional analysis, higher levels of objectively-measured walking were associated with larger hippocampal volume in females only^21^, with a follow-up study indicating this effect was confined to the subiculum^39^. It is important to note that only 26 males were included in this study, limiting the statistical power required to detect an effect. Although we detected sex as a moderator of the relationship between physical activity and brain volume in our large sample, this work requires further confirmation utilising longitudinal analyses and exercise interventions to further understand the nature of these sex differences.

Our study did not detect a moderating effect of the *BDNF* Val66Met polymorphism nor the *APOE* ε4 allele on the physical activity and brain volume relationship. This is in contrast to previous work by our group: we identified a link between physical activity and hippocampal volume in *BDNF* Val/Val homozygotes, but not Met carriers^26^. In addition, other studies have identified moderating effects of the *BDNF* Val66Met polymorphism in the relationship between physical activity and episodic memory (hippocampal-dependent function)^40, 41^. In regards to the potential moderating effects of the *APOE* ε4 allele, a previous cross-sectional evaluation of cardiorespiratory fitness and brain volume identified no effect of genotype^42^. Yet, over an 18 month follow up, Smith and colleagues identified higher rates of hippocampal atrophy in ε4 carriers that were low active, yet stable hippocampal volume was observed in individuals that were high active^4^. In the current analysis, our large sample ensured we were sufficiently powered to detect moderating effects of genotype; thus, our divergent findings could be attributed to demographic differences of the studied cohorts, or other methodological differences across the studies. Furthermore, it is likely that a myriad of genetic factors influence the relationship between physical activity and the brain. Thus, investigations of combinations of genetic factors, such as in polygenic risk scores, as moderators of the physical activity-brain volume relationship warrant investigation, particularly in large samples.

Our study had some limitations. The causal direction of our findings must be interpreted with caution. More specifically, it is possible that in males only, brain volume may play a role in influencing participation in physical activity. Future longitudinal and interventional research may aid in elucidating the nature of the moderating effect of sex on the relationship between physical activity and brain volume. Our study investigated sex as a moderator of the relationship between physical activity and brain volume; however, our research is unable to identify *gender* differences (referring to social, environmental and behavioural factors), which may also play a role in the differential effects of physical activity on brain health. Finally, accelerometer data and MRI brain imaging were not conducted at the same timepoint, and thus variation in the time difference between these measures may have influenced our findings. Nevertheless, for privacy issues, and to avoid self-identification, dates of these assessments were not available in order to calculate time between assessments.

Our findings using a large dataset provides further evidence that habitual physical activity levels are associated with larger brain volume. In addition, we found that only men received the benefit in terms of larger total grey matter and right hippocampal volumes. Future research is needed to both understand whether there is variability in response to exercise interventions between the sexes and the potential moderating effect of other genetic factors, in order to design individualised intervention programmes.

## Supporting information

eFigure 1

eTable 1

## Data Availability

All data used in this study can be accessed through application to the UK Biobank.

## Acknowledgements

The data used to generate this work were obtained from the UK Biobank (application 45567). We are grateful to all UK Biobank study participants, who generously donated their time and the UK Biobank team for their work collecting and processing the data. The authors acknowledge Kelsey Sewell (Murdoch University) for assistance with figure preparation.

## Notes

### Competing Interest Statement

The authors have declared no competing interest.

### Funding Statement

This study did not receive any funding.

### Author Declarations

Ethical approval was provided by the NHS National Research Ethics Committee (11/NW/0382) for the UK Biobank.

