## Supplementary material for "Non-modifiable factors as moderators of the relationship between physical activity and brain volume: A cross-sectional UK Biobank study": eFigure 1

***eFigure 1.*** *Flow chart of participant numbers for UK Biobank objectively-measured physical activity and volumetric magnetic resonance imaging analysis*

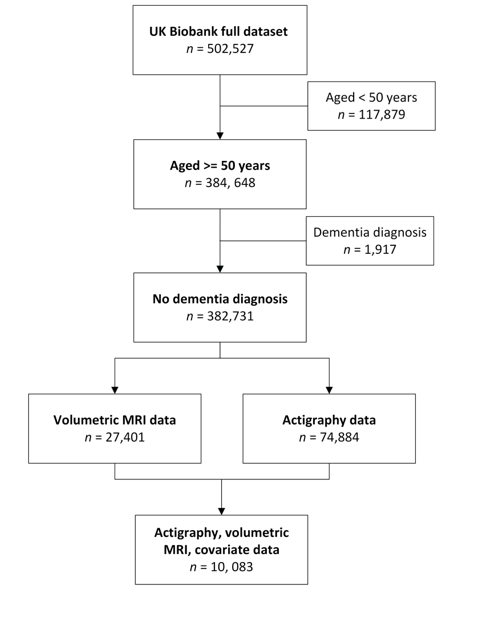
