## Supplementary material for "Non-modifiable factors as moderators of the relationship between physical activity and brain volume: A cross-sectional UK Biobank study": eTable 1

***eTable 1:*** *Descriptive statistics of UK Biobank participants at baseline (2006 - 2010) aged over 50 years with accelerometer and MRI data, stratified by apolipoprotein* ε*4 allele carriage and brain-derived neurotrophic factor Val66Met single nucleotide polymorphism carriage.*

|  | ***APOE* ε4 non-carriers**  (*n* = 7329) | ***APOE* ε4 carriers**  (*n* = 2754) | **Test statistic** | ***BDNF* Val/Val**  (*n* = 6609) | ***BDNF* Met carriers**  (*n* = 3474) | **Test statistic** |
| --- | --- | --- | --- | --- | --- | --- |
| **Age (years)** | 58.8 ± 5.1 | 58.5 ± 5.0 | *t* = 2.32* | 58.7 ± 5.1 | 58.7 ± 5.1 | *t* = 0.5 |
| **Body mass index** | 26.5 ± 4.1 | 26.4 ± 4.1 | *t* = 1.1 | 26.5 ± 4.1 | 26.4 ± 4.2 | *t* = 1.1 |
| **Physical activity (accelerometer quantified daily accelerations)** | 27.5 ± 8.7 | 27.7 ± 8.7 | *t* = 1.0 | 27.5 ± 8.7 | 27.6 ± 8.6 | *t* = 0.7 |
| **Accelerometer wear time (days)** | 6.4 ± 1.4 | 6.4 ± 1.4 | *t* = 0.7 | 6.4 ± 1.4 | 6.4 ± 1.3 | *t* = 0.5 |
| **Education** | | | | | | |
| College or University degree, % (*n*) | 45.7 (3348) | 45.8 (1262) | χ^2^ = 11.7 | 45.7 (3021) | 45.7 (1589) | χ^2^ = 3.4 |
| A levels/AS levels or equivalent, % (*n*) | 12.8 (936) | 13.0 (357) |  | 12.7 (838) | 13.1 (455) |  |
| O levels/DCSEs or equivalent, % (*n*) | 18.9 (1388) | 19.9 (548) |  | 19.2 (1267) | 19.3 (669) |  |
| CSEs or equivalent, % (*n*) | 3.3 (243) | 2.8 (77) |  | 3.1 (205) | 3.3 (115) |  |
| NVQ/HND/HNC or equivalent, % (*n*) | 5.3 (390) | 6.2 (171) |  | 5.8 (383) | 5.1 (178) |  |
| Other professional qualification, % (*n*) | 6.3 (460) | 5.2 (144) |  | 5.9 (392) | 6.1 (212) |  |
| None of the above, % (*n*) | 7.7 (564) | 7.1 (195) |  | 7.7 (503) | 7.4 (256) |  |
| **Sex, female % (*n*)** | 53.3 (3903) | 54.1 (1493) | χ^2^ = 0.7 | 53.4 (3529) | 53.7 (1867) | χ^2^ = 0.1 |
| **Major depression % (*n*)** | 21.3 (1564) | 21.6 (596) | χ^2^ = 0.1 | 21.8 (1443) | 20.6 (717) | χ^2^ = 1.9 |
| **Townsend Deprivation Index** | -2.1 ± 2.6 | -2.0 ± 2.7 | *t =* 1.0 | -2.0 ± 2.6 | -2.0 ± 2.6 | *t* = 0.1 |
| **Overall health rating** | | | | | | |
| Excellent, % (*n*) | 25.6 (1879) | 26.3 (725) | χ^2^ = 4.3 | 25.8 (1703) | 25.9 (901) | χ^2^ = 0.7 |
| Good, % (*n*) | 60.4 (4426) | 60.4 (1662) |  | 60.4 (3990) | 60.4 (2098) |  |
| Fair, % (*n*) | 12.4 (911) | 12.1 (332) |  | 12.4 (822) | 12.1 (421) |  |
| Poor, % (*n*) | 1.4 (104) | 1.3 (35) |  | 1.3 (89) | 1.4 (50) |  |
| **Vascular problems % (n)** | 14.3 (1044) | 14.2 (390) | χ^2^ = 5.5 | 14.5 (958) | 13.8 (476) | χ^2^ = 3.9 |
| **Diabetes % (n)** | 2.6 (190) | 2.2 (61) | χ^2^ = 1.5 | 2.4 (160) | 2.6 (91) | χ^2^ = 0.9 |
| **Head scaling factor** | 1.3 ± 0.1 | 1.3 ± 0.1 | *t =* 0.2 | 1.3 ± 0.1 | 1.3± 0.1 | *t =* 0.7 |
| **Cortical grey matter (normalised; cm^3^)** | 606.4 ± 36.8 | 606.8 ± 37.0 | *t* = 0.5 | 606.5 ± 37.1 | 606.6 ± 36.5 | *t* = 0.2 |
| **Grey matter (normalised; cm^3^)** | 778.9 ± 43.1 | 779.9 ± 43.2 | *t* = 0.9 | 779.2 ± 43.4 | 779.2 ± 42.8 | *t* = 0.01 |
| **Left hippocampal volume (normalised; mm^3^)** | 3748 ± 452 | 3720 ± 455 | *t =* 2.7 | 3736 ± 454 | 3749 ± 452 | *t* = 1.3 |
| **Right hippocampal volume (normalised; mm^3^)** | 3854 ± 463 | 3830 ± 470 | *t =* 2.3 | 3850 ± 462 | 3844 ± 471 | *t* = 0.6 |

**p* < 0.05 ; Chi-square analyses for categorical variables and independent sample t-tests were conducted for continuous variables. Abbreviations: *APOE*, apolipoprotein E; *BDNF* Val66Met, brain-derived neurotrophic factor Valine66Methionine single nucleotide polymorphism.
